# SARS-CoV-2 Antibody persistence in COVID-19 convalescent plasma donors

**DOI:** 10.1101/2021.03.24.21254260

**Authors:** Clara Di Germanio, Graham Simmons, Kathleen Kelly, Rachel Martinelli, Orsolya Darst, Mahzad Azimpouran, Mars Stone, Kelsey Hazegh, Eduard Grebe, Shuting Zhang, Peijun Ma, Marek Orzechowski, James E Gomez, Jonathan Livny, Deborah T. Hung, Ralph Vassallo, Michael P. Busch, Larry J. Dumont

**Affiliations:** Vitalant Research Institute, San Francisco, CA, USA; Department of Laboratory Medicine, University of California San Francisco, San Francisco, CA, USA; Vitalant Research Institute, Denver, CO, USA; Vitalant, Scottsdale, Arizona, USA; University of Colorado School of Medicine, Aurora, CO, USA; Infectious Disease and Microbiome Program, Broad Institute of MIT & Harvard, 415 Main Street, Cambridge, MA, 02142, USA; Department of Molecular Biology and Center for Computational and Integrative Biology, Massachusetts General Hospital, 185 Cambridge Street, Boston, MA, 02114, USA; Department of Genetics, Harvard Medical School, 77 Avenue Louis Pasteur, Boston, MA, 02115, USA

**Keywords:** SARS-CoV-2 Antibody, Antibody waning, convalescent plasma

## Abstract

**Background:** Antibody response duration following SARS-CoV-2 infection tends to be variable and depends on severity of disease and method of detection.

**Study design and methods:** COVID-19 convalescent plasma (CCP) from 18 donors was collected longitudinally for a maximum of 63 - 129 days following resolution of symptoms. All the samples were initially screened by the Ortho Total Ig test to confirm positivity and subsequently tested with 7 additional direct sandwich or indirect binding assays (Ortho, Roche, Abbott, Broad Institute) directed against a variety of antigen targets (S1, RBD, and NC), along with 2 neutralization assays (Broad Institute live virus PRNT and Vitalant Research Institute Pseudovirus RVPN).

**Results:** The direct detection assays (Ortho Total Ig total and Roche Total Ig) showed increasing levels of antibodies over the time period, in contrast to the indirect IgG assays that showed a decline.

Neutralization assays also demonstrated declining responses; the VRI RVPN pseudovirus had a greater rate of decline than the Broad PRNT live virus assay.

**Discussion:** These data show that in addition to variable individual responses and associations with disease severity, the detection assay chosen contributes to the heterogeneous results in antibody stability over time. Depending on the scope of the research, one assay may be preferable over another. For serosurveillance studies, direct, double Ag-sandwich assays appear to be the best choice due to their stability; in particular, algorithms that include both S1 and NC based assays can help reduce the rate of false-positivity and discriminate between natural infection and vaccine-derived seroreactivity.

## INTRODUCTION

In the past several months, research on Coronavirus Disease 2019 (COVID-19) immune response has confirmed that the majority of infected individuals mount antibody responses to the severe acute respiratory syndrome coronavirus 2 (SARS-CoV-2), the virus that causes the disease.^1^ Increasing evidence suggests that more rapid and potent humoral responses correlate with the severity of disease,^2, 3^ likely due to greater and longer exposure to the viral antigen. The antibody response to acute viral infections typically declines from peak titers following recovery from acute infection. Conflicting reports have described the rapidity of antibody decline following SARS-CoV-2 infection, with some studies reporting a rapid decay of anti-SARS-CoV-2 antibodies,^3,4,5,6^ while others have described stable short-term responses^7^ or even a sustained response, up to 8 months after infection.^8^ In keeping with the strength of the initial humoral response correlating with severity of disease, many of the studies demonstrating rapid waning of anti-viral responses have been performed in asymptomatic individuals or patients with mild symptoms. Indeed, in a direct comparison, antibodies decayed more rapidly in mild cases compared to severe infection.^9^ This finding is in line with previous studies for other coronaviruses.^10^ However, there is also evidence that some of the variability seen in these studies is due to the serological assays deployed.^8^ Different antibody isotypes, antibodies targeting different antigens and epitopes, and different assay formats (indirect, direct), are likely to wane with differing kinetics, making some immunoassay approaches more effective than others. The most common commercially available assays target either the spike subunit (S1) that mediates viral entry, the receptor binding domain (RBD) of S1 which binds to its human cellular receptor angiotensin-converting enzyme 2 (ACE2), or the nucleocapsid (NC) protein that encapsulates the viral genome.^11^

Using 18 repeat donors of COVID-19 convalescent plasma (CCP) we studied the kinetics of antibody evolution and decline up to 129 days after resolution of COVID-19 symptoms. Eight antibody binding assays on different platforms and targeting different antibody isotypes and viral antigens were investigated, as well as two assays of viral neutralization (Table 1). This allowed in-depth comparison between assays and antigen targets in order to identify assays that may be suited for different purposes such as serodiagnosis of recently acquired infection, serosurveillance, correlates of immune protection, or potency of CCP. Assays that effectively detect and quantify long-lasting serological responses are important tools for making accurate seroprevalence estimates of previous infection to track incidence over time. Alternately, immunoassays that demonstrate a rapidly waning response may be useful for performing recency studies, identifying hotspots of infection and predicting subsequent protective immunity at the individual and population level (i.e., herd immunity). High-throughput assays waning in parallel with neutralizing antibody response may also be particularly valuable in the characterization of CCP neutralizing antibody content.

**Table 1.**
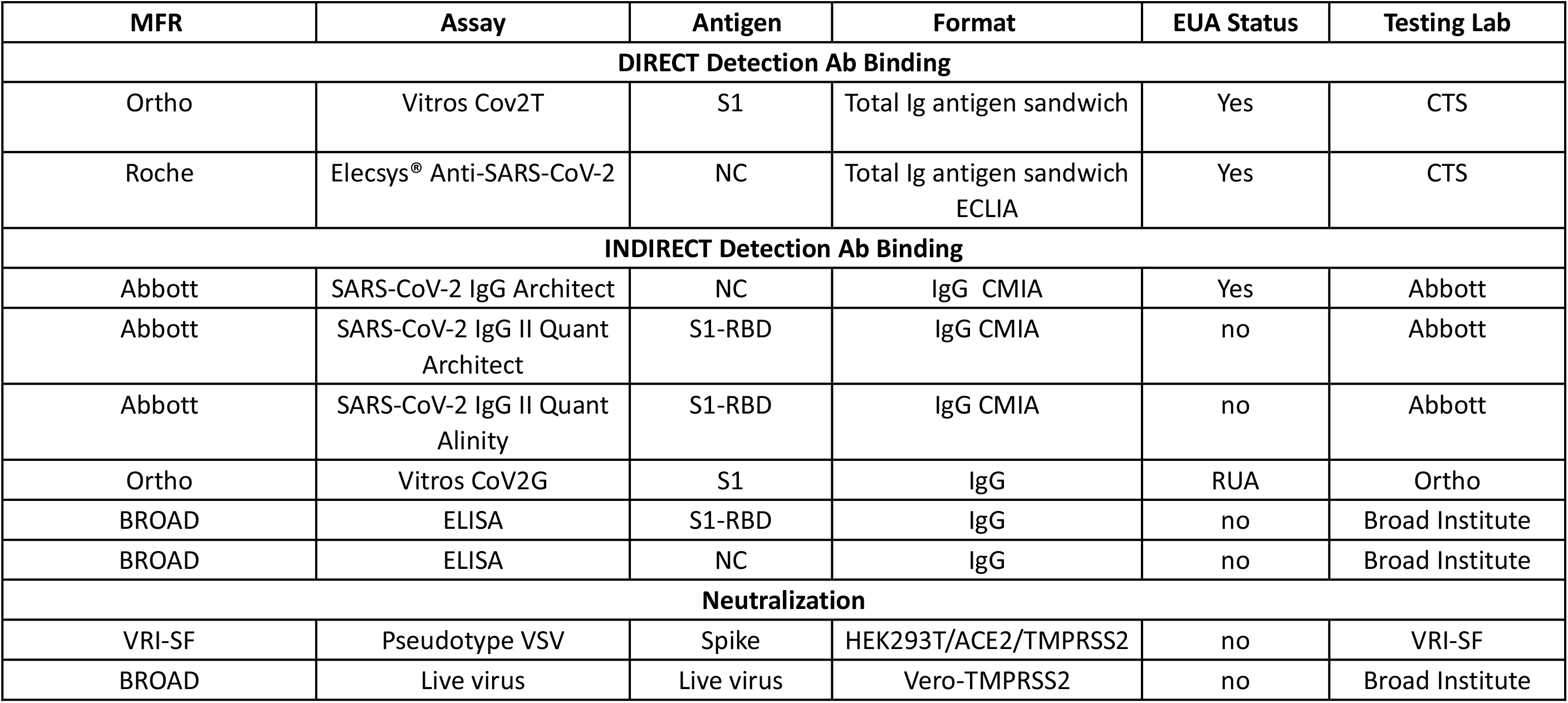
SARS-CoV-2 Antibody Binding and Neutralization Assays.

## METHODS

### COVID-19 Convalescent Plasma

CCPs were collected in the Vitalant system following FDA Guidance for donor eligibility^12^ as previously described.^13^ These criteria evolved throughout the study period due to testing availability and evolution of the pandemic in the United States. Evidence of COVID-19 was required in the form of a documented positive SARS-CoV-2 molecular or serologic test, and complete resolution of symptoms initially at least 14 days prior to donation (with a negative molecular test if <28 days), but then a minimum of 28 days was implemented. All CCP donors were also required to meet traditional allogeneic blood donor criteria. At the time of plasma collection, donors consented to use of de-identified donor information and test results for research purposes. Medical Director approval was obtained for CCP collection every 7 days for interested donors. Between April 8 and October 20, 2020, approximately 50,000 units were released from 7976 unique donors. All CCPs were tested for SARS-CoV-2 total Ig antibody using the Ortho VITROS CoV2T assay at our central testing laboratory (Creative Testing Solutions [CTS], Scottsdale, AZ). CCP qualification requires the signal-to-cutoff ratio S/CO of this test to be at least 1.0. At least 2 collections were conducted for 2507 donors in this period. Of these, 275 had greater than four donations and an interval from the first to last donation of over 60 days. We selected a subset of 19 unique donors for evaluation of the persistence of SARS-CoV-2 Ab with date range from first to last donation 47-99 days (resolution of symptoms to last donation 63-129 days) and 4-12 total donations. One donor was subsequently removed from the analysis set for cause because only 2 retained serum samples were retrievable and these were negative for neutralizing antibody. For the present analysis, donor COVID-19 symptom and SARS-CoV-2 testing histories were reviewed. The time course of longitudinal donations was determined based on the date of resolution of all disease symptoms.

## Antibody measurement

### Direct Detection

#### Ortho VITROS Anti-SARS-CoV-2 Total

The Ortho VITROS Anti-SARS-CoV-2 Total (CoV2T, Ortho-Clinical Diagnostics, Inc., Rochester, NY) was used at CTS to detect total (IgG, IgM and IgA) antibodies against the spike S1 protein, as previously described.^13^ Briefly, serum samples are quickly vortexed, loaded on Ortho VITROS XT-7200 or 3600 instruments (Ortho Clinical Diagnostics, Raritan, NJ) and programmed for the CoV2T test following the manufacturer’s instructions.^14^ The S1 antigens coated on the assay wells bind S1 antibodies from human serum which, in turn, bind to a secondary HRP-labeled S1 antigen in the conjugate reagent forming a sandwich. The addition of signal reagent containing luminol generates a chemiluminescence reaction that is measured by the system and quantified as the ratio of the signal relative to the cut-off value generated during calibration. An S/CO ≥1 is considered positive.

#### Roche cobas Anti-SARS-CoV-2

The Elecsys Anti-SARS-CoV-2 immunoassay (Roche NC) was run at CTS on the cobas e441 analyzer (Roche Diagnostics, Indianapolis, IN) to detect antibodies against the SARS-CoV-2 nucleocapsid protein. Plasma samples are initially incubated with biotinylated and ruthenium-labelled SARS-CoV-2 recombinant nucleocapsid antigens and any antibody present in the solution is sandwiched between the two. Subsequently, streptavidin-coated microparticles are added to the mixture to bind the biotin. The magnetic particles drive the complexes to the electrode, where a chemiluminescent signal is emitted and measured as the ratio between the signal and the cut-off obtained during calibration. Similarly to the VITROS, a S/CO ≥1 is considered positive.^15^

### Indirect detection

#### Ortho VITROS Anti-SARS-CoV-2 IgG

Levels of IgG antibodies were measured in plasma by the Ortho VITROS Anti-SARS-CoV-2 IgG (CoV2G, Ortho-Clinical Diagnostics, Inc., Rochester, NY) at Ortho Diagnostics using a quantitative RUA assay. Similar to the VITROS CoV2T, in the first step the antibodies present in the specimen bind to the S1 spike on the testing wells. However, in the following stage, HRP-conjugated murine monoclonal anti-human IgG antibodies are added, targeting the antibody portion of the complex. When the luminogenic substrate is added, chemiluminescence is then measured and quantified as a S/CO, with values above 1 considered positive.^16^

#### Abbott Architect SARS-CoV-2 IgG

A qualitative SARS-CoV-2 IgG assay targeting IgG against nucleocapsid protein was performed by Abbott (Architect NC IgG, Abbott Diagnostics, Abbott Park, IL) on the Architect platform using chemiluminescent microparticle immunoassay (CMIA) technology.^17^ An additional quantitative assay, SARS-CoV-2 IgG II Quant (not available in the U.S.) was performed on the Architect and Alinity platforms to detect and measure IgG against the RBD of the S1 protein of the virus. Plasma samples are incubated with SARS-CoV-2 antigen coated on magnetic microparticles which bind IgG antibodies against SARS-CoV-2. These complexes are then incubated with anti-human IgG acridinium-labeled conjugate, which result in a chemiluminescence reaction upon addition of trigger solution. Results are reported as an index value (S/C) comparing RLU (relative light units) from the samples and the calibrator for the IgG assay and as Arbitrary Units/mL (AU/mL) comparing RLU from the sample relative to the RLUs obtained from a 6-point calibration curve for the IgG II assay. Values ≥1.40 S/C on the IgG assay and ≥ 50 AU/mL on the IgG II assay are considered positive.

#### Broad Institute ELISA

Quantitative ELISAs to measure antibodies to the receptor binding domain (RBD) and nucleocapsid proteins (BROAD RBD and BROAD NC, respectively) were developed and performed at the Broad Institute (Cambridge, MA). 50 μL of 1:100 diluted serum samples were added to MaxiSorp 384-well microplates (Sigma) pre-coated with 50 μL/well of 2,500 ng/ml of SARS-CoV-2 RBD and incubated for 30 min at 37 °C. Plates were then washed and then 50 μL/well of 1:25,000 diluted detection antibody solution (HRP-anti human IgG and IgM, Bethyl Laboratory) was added. After an incubation for 30 min at RT, plates were washed and 40 μL/well of Pierce TMB peroxidase substrate (ThermoFisher) was added. The reaction was stopped by adding 40 μL/well of stop solution (0.5 M H2SO4). The OD was read at 450 nm and 570 nm on a BioTek Synergy HT. For control antibodies CR3022 IgG1 and IgM (Absolute Antibody) dilution curves, the antibodies were diluted to a concentration of 1μg/mL in dilution buffer and duplicate 12 two-fold serial dilution curves were generated. Sample concentrations were estimated based on the standard curve.

### Neutralization assays

#### Broad Institute Live Virus Neutralization

Live-virus SARS-CoV-2 antibody neutralization was performed at Broad Institute on a high throughput platform (BROAD PRNT). Vero E6-TMPRSS2 were seeded at 10,000 cells per well the day prior to infection in a CellCarrier-384 ultra-microplate (Perkin Elmer). Patient serum samples were tested at a starting dilution of 1:40 and were serially diluted 2-fold up to eight dilution spots. Serially diluted patient sera were mixed separately with diluted SARS-CoV-2 live virus (D614) and incubated at 37°C with 5% CO2 for 1 hour; after which the sera-virus complexes were added to the Vero E6-TMPRSS2 cells and incubated at 37°C with 5% CO2 for 48 hours. Cells were then fixed using 4% paraformaldehyde in PBS for 2 hours at room temperature, washed, and incubated with diluted anti-SARS-CoV/SARS-CoV-2 nucleoprotein mouse antibody (Sino Biological) for 1.5 hours at room temperature. They were subsequently incubated with Alexa488-conjugated goat anti-mouse (Jackson ImmunoResearch Labs) for 45 mins at room temperature, followed by nuclear staining with Hoechst 33342 (Thermo Fisher Scientific). Fluorescence imaging was performed using the Opera Phenix™ High Content Screening System (Perkin Elmer). Half-maximal inhibitory dilutions (ID50) were determined using a four-parameter, nonlinear curve fitting algorithm. Samples whose curves lay above 0.5 for all the data points were considered non-neutralizing, with ID_50_=20, while samples whose curves fell below 0.5 were considered highly neutralizing and assigned an ID_50_=10,240.

#### Vitalant Research Institute (VRI) Pseudovirus Neutralization

Serum samples were tested at VRI for SARS-CoV-2 reporter viral particle neutralization (RVPN) as previously described^13, 18, 19^ using the Wuhan-Hu-1 spike sequence (GenBank: MN908947.3) modified by addition of the D614G mutation and removal of 21 C-terminal amino acids demonstrated to enhance incorporation into viral particles. Recombinant vesicular stomatitis virus (VSV) containing firefly luciferase gene (Kerafast, Boston, MA) and incorporating SARS-CoV-2 spike were added to heat inactivated samples diluted four-fold, together with positive, negative and no-serum controls. The resulting mix was incubated and then added to 96-well plates containing ACE2 and TMPRSS2 expressing HEK293T cells. Eighteen to 24 hours later, luciferase activity was measured on a chemiluminescence reader (BMG CLARIOStar, BMG LABTECH Inc., Cary, NC) after lysing the cells. Neutralization titers were calculated as a percentage of no-serum control and the NT_50_ was estimated from the dilution curve using Prism8 (GraphPad Software, San Diego, CA). Titers below 40 were considered non-neutralizing.

### Statistical Analysis

Primary assay outcomes from VRI RVPN, BROAD PRNT, BROAD NC, BROAD RBD, Ortho CoV2T, Abbott Alinity S IgG, and Abbott Architect S IgG were logarithmically (base 10) transformed to meet the regression model assumption requirements. The Roche NC, Ortho CoV2G, and Abbott Architect NC IgG outcome signals remained in the reported scale. These data were then standardized using a z-transformation by assay:

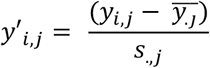

Standardized outcomes were fit to a mixed linear model of z-signal regressed on the time from resolution of signs and symptoms as reported by the donor. Random variables of intercept and slope were nested within assay with repeated measures for serum sample (DIN) and donor as subject. Thus, the overall slope and intercept solution for each assay was adjusted by individual donor random effects. Plots of the regression solution for each assay were constructed showing the overall estimated z-signal over time with the empirical best linear unbiased predictor (EBLUP) for each donor. Pearson conditional residuals were examined to evaluate the final model assumptions and fit; and were found to support the key assumptions of independence and normality (Proc Stdize and Proc Mixed, SAS v9.4, SAS Institute, Cary, NC). Hypotheses tests were conducted using the model at p>0.05 as significant. P-values were not adjusted for multiple comparisons.

To determine if the sample selected was representative of the total number of donors with multiple donations over the period, we evaluated the Ortho VITROS CoV2T signal over time from first donation for the 275 donors with >60d maximum observation period at >4 donations. These data were evaluated by regression as described above and compared to the subset of 18 subjects chosen for this study.

## RESULTS

Figure 1 shows the Ortho CoV2T S/CO results for 275 unique donors with at least 4 donations spanning a minimum of 60 days during the observation period. The 18 unique donors selected for additional antibody testing in this study are highlighted, and the regression solutions for all donors and the selected donor subset are shown. The 18 (14 male / 4 female) evaluable donors all had symptomatic COVID-19 (median 17 days of symptoms; range 3-22). Seventeen were confirmed by positive swab testing, the other by antibody testing. The donors’ median age was 57 (range 22-73). Evaluable serum samples were available for testing through a maximum of 63-129 days following the resolution of COVID-19 signs and symptoms.

**Figure 1.**
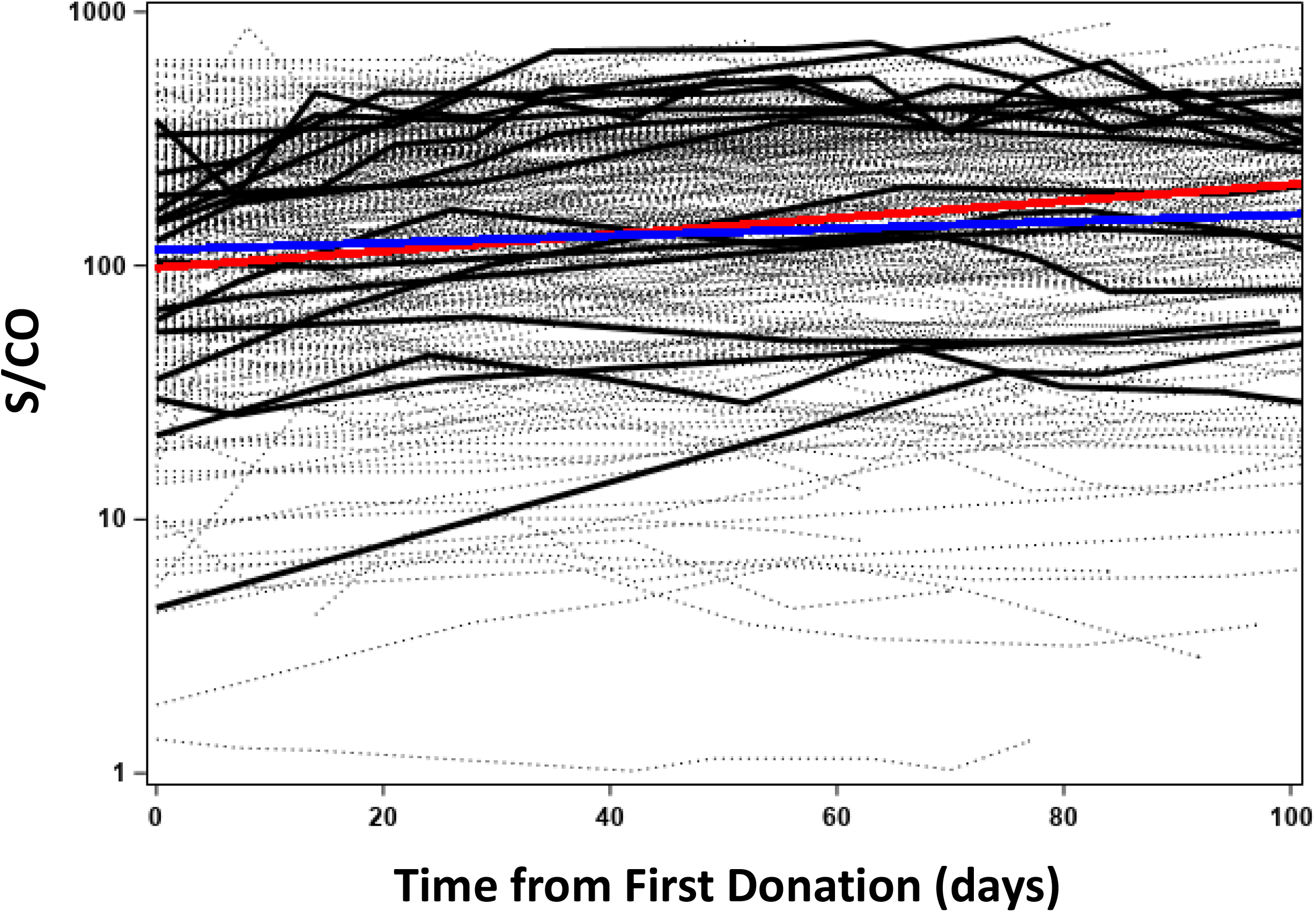
Study Sample Set is Representative of All Repeated Donors. Ortho CoV2T (Total Ig) S/CO signal for unique donors with at least 4 donations and greater than 60 days from the last donation to the first CCP donation 8 April – 20 October 2020. Light dashed lines - 275 repeated donors Blue line – overall regression solution for 275 donors Heavy black lines – 18 donors selected for present study Red line – overall regression solution for 18 donors

The 18 donors had 873 evaluable observations. The reported, non-transformed assay signals by donor for each assay are shown in Figure 2. Large differences are noted between donor antibody assay signals in each assay. Figure 3 displays the regression result for the standardized outcomes for each assay showing the individual solutions for each donor and the overall average. The standardized slopes for each assay are shown in Table 2 and Figure 4. Total antibody levels determined by direct-detection, sandwich methods (Ortho CoV2T, Roche NC) increased over the course of observation. The rate of change over time (slopes) were not different between Ortho CoV2T and Roche NC, p=0.66. The 6 indirect, IgG assays showed declining levels. The declines in the standardized assay outcome (slope) for the indirect binding IgG assays comprised of different S1, RBD and NC antigens were not significantly different, p=0.70. The BROAD PRNT assay values declined throughout the observation period at a rate not different than the indirect binding assays (p=0.75). The VRI RVPN pseudovirus test demonstrated consistent decline (−0.0230 ± 0.0018 95%CI: -0.0266 to -0.0194, p<.0001), and was significantly greater than the BROAD PRNT waning, p< 0.0001.

**Table 2:** Change in Antibody Signal over Time – standardized units per day (slope ± SE; 95% confidence interval)

| **Assay** | **Slope** | **95% CI** | **p** |
| --- | --- | --- | --- |
| Ortho VITROS CoV2T | 0.0066 ± 0.0017 | 0.0032 - 0.0099 | <.0001 |
| Roche NC | 0.0052 ± 0.0017 | 0.0018 - 0.0086 | 0.003 |
| Ortho VITROS CoV2G | -0.0059 ± 0.0017 | -0.0093 - -0.0026 | <.0001 |
| BROAD PRNT ID<sub>50</sub> | -0.0065 ± 0.0018 | -0.0101 - -0.0030 | <.0001 |
| BROAD RBD ELISA | -0.0066 ± 0.0018 | -0.0102 - -0.0031 | <.0001 |
| BROAD NC ELISA | -0.0072 ± 0.0018 | -0.0108 - -0.0037 | <.0001 |
| Abbott Architect-S IgG II Quant | -0.0087 ± 0.0017 | -0.0120 - -0.0053 | <.0001 |
| Abbott Alinity-S IgG II Quant | -0.0088 ± 0.0017 | -0.0121 - -0.0054 | <.0001 |
| Abbott Architect-NC IgG | -0.0093 ± 0.0017 | -0.0126 - -0.0059 | <.0001 |
| VRI RVPN NT<sub>50</sub> | -0.0230 ± 0.0018 | -0.0266 - -0.0194 | <.0001 |

**Figure 2.**
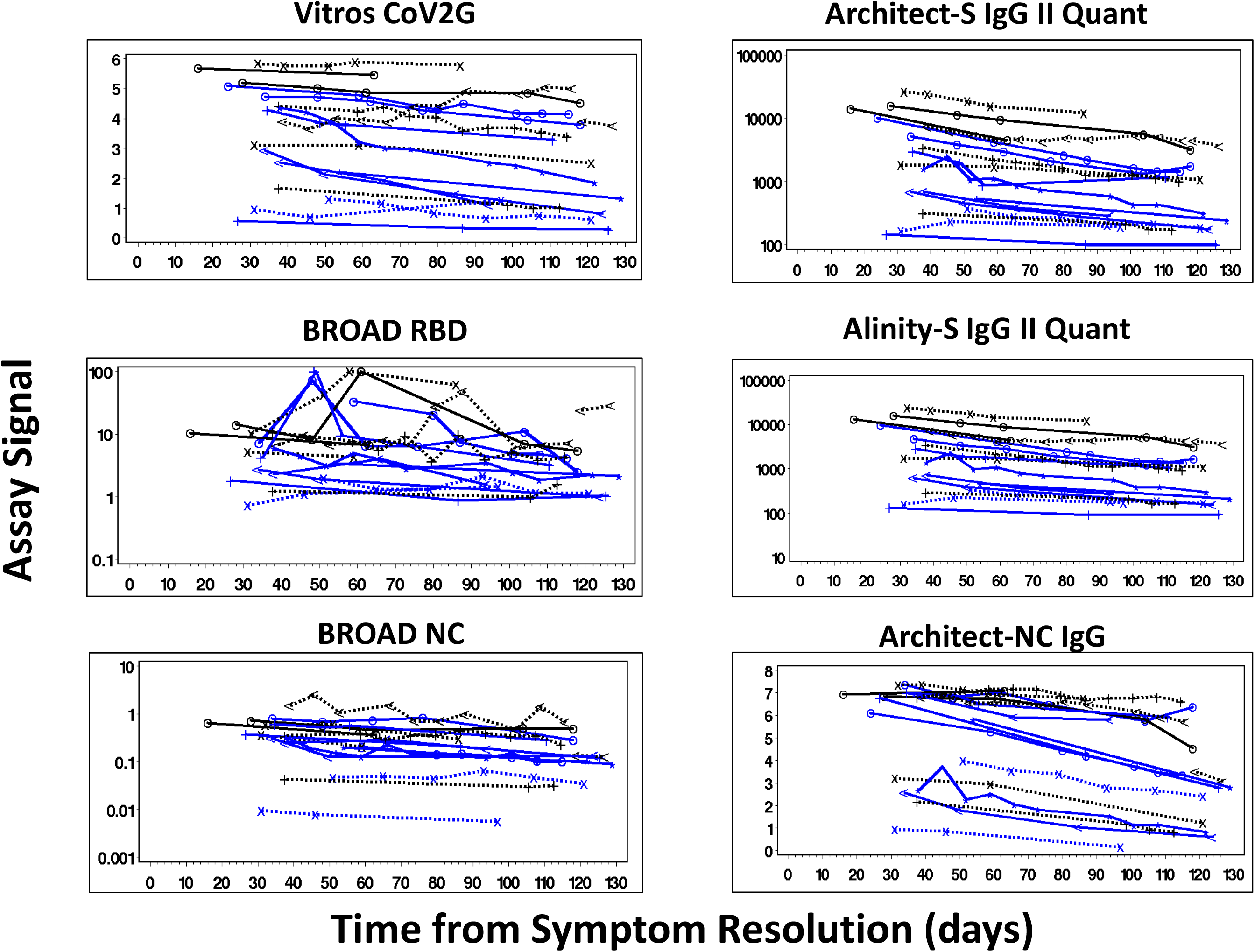
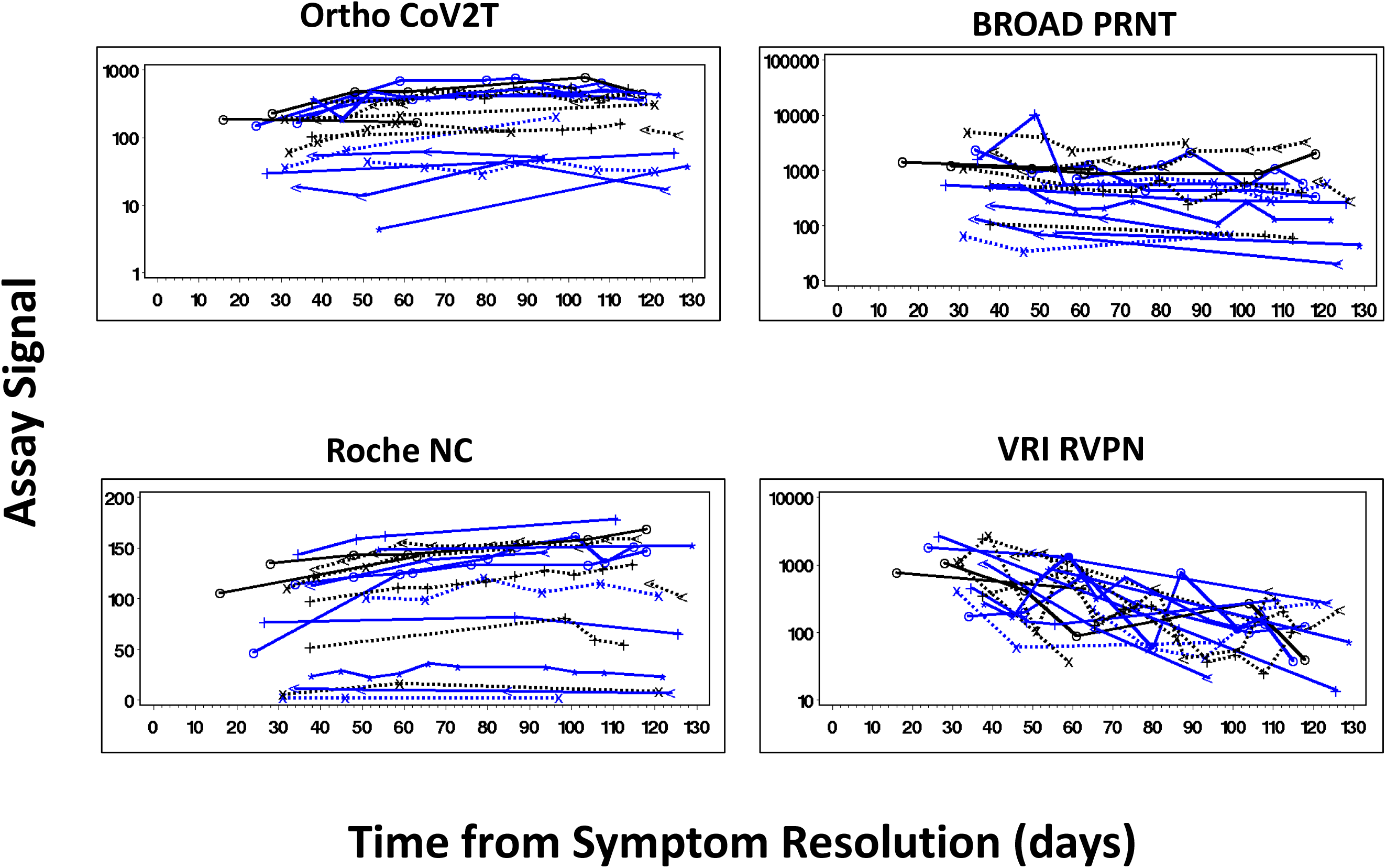
Antibody Stability Over Time Post Resolution of COVID-19 Symptoms – Assay Signals as Reported. See Table 1 for assay descriptions. 2A: Indirect detection method assays using anti-IgG secondary antibodies 2B: Left panels - Direct detection method assays using SARS-CoV-2 antigen detection Right panels – neutralizing antibody assays

**Figure 3.**
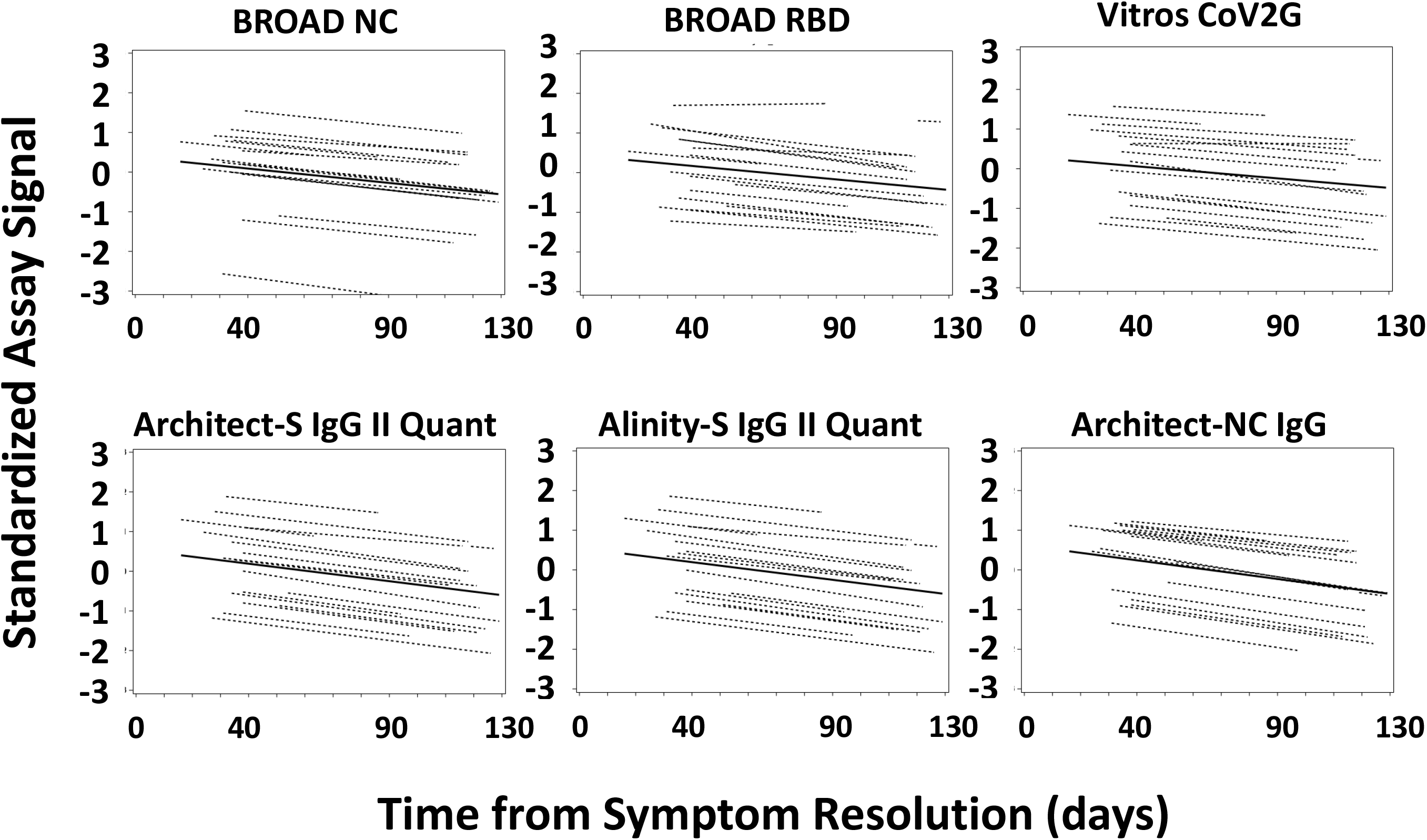
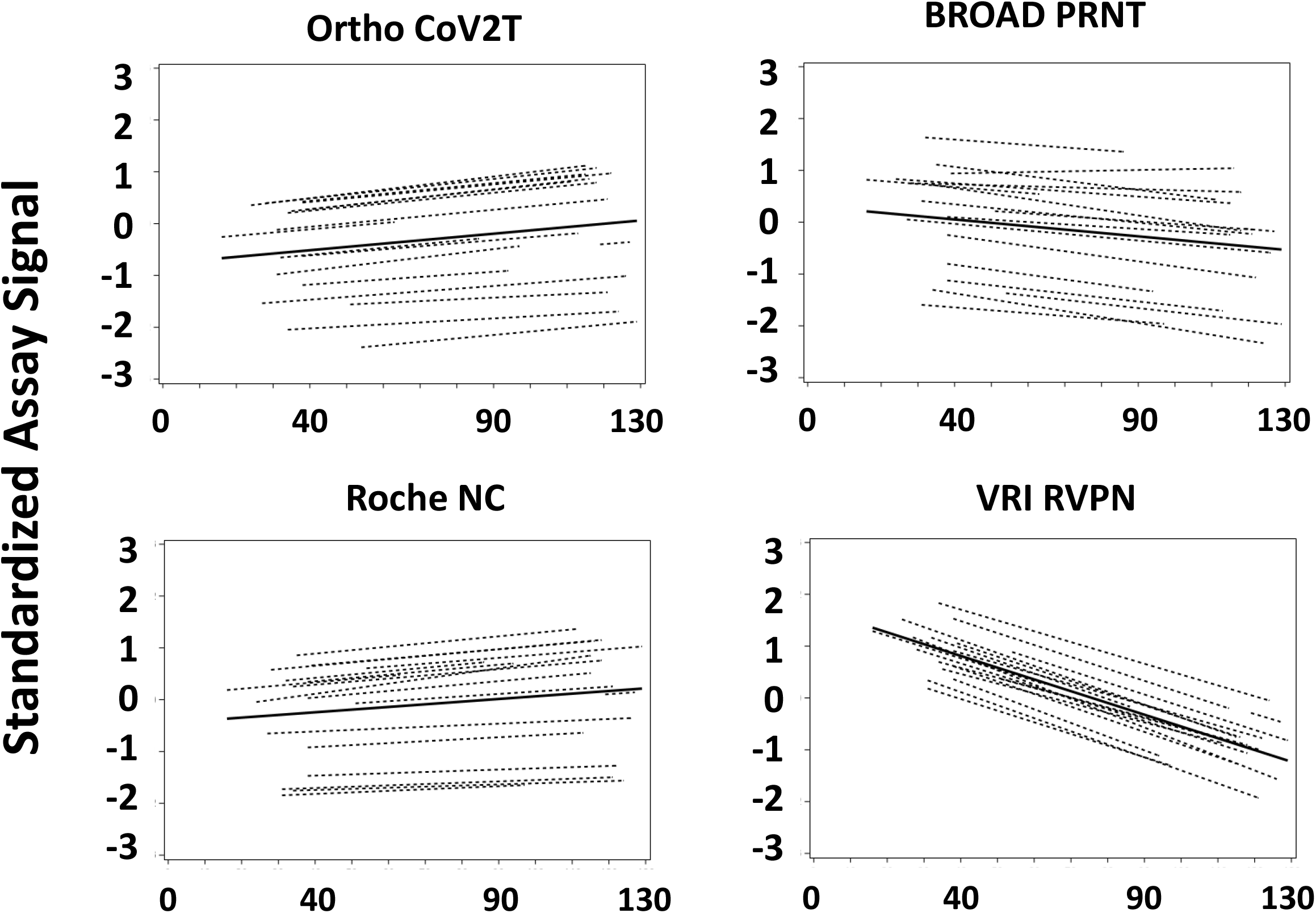
Antibody Stability Over Time Post Resolution of COVID-19 Symptoms – Standardized Assay Signals. See Table 1 for assay descriptions. 2A: Indirect detection method assays using anti-IgG secondary antibodies 2B: Left panels - Direct detection method assays using SARS-CoV-2 antigen detection Right panels – neutralizing antibody assays

**Figure 4.**
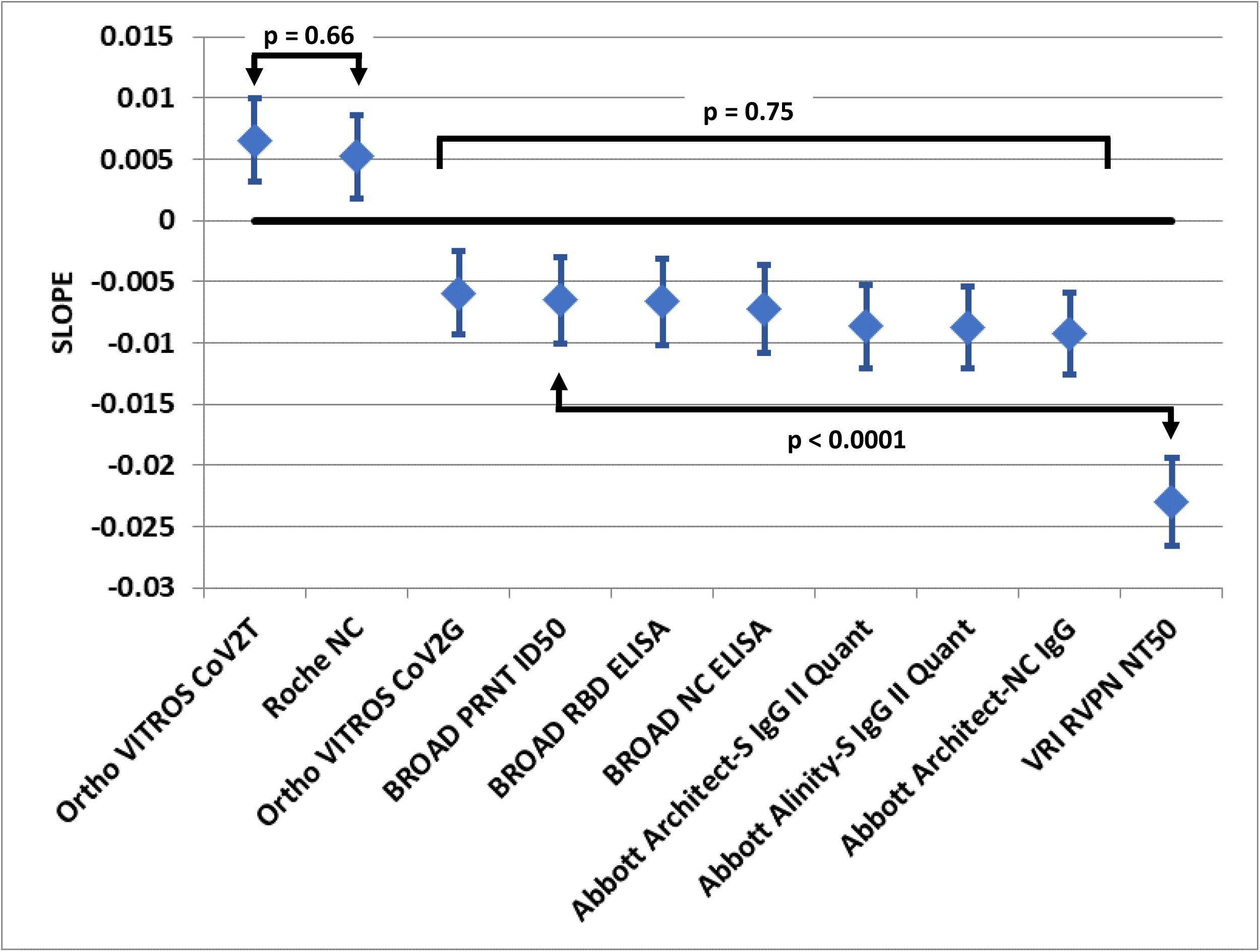
Regression Solution Standardized Slope of Signal Decay Over Time –. standardized units per day. Diamond=estimate, error bars 95% CI. No difference is observed for direction detection methods. Indirection detection methods slopes are not different in a global hypothesis test. The two neutralization assays have significantly different rates of decline.

## DISCUSSION

This study accessed longitudinal samples from CCP donors with intervals between resolution of disease symptoms and last donation of 63-129 days and 3-12 total donations per donor to evaluate and compare the persistence of antibody reactivity of 10 SARS-CoV-2 antibody assays. These assays included six commercial assays with different antigen targets (S1, RBD, NC) and detection formats (direct detection by antigen labeled conjugate; indirect detection using anti-human IgG labeled conjugate), S1 RBD and NC-based assays developed at the Broad Institute, and live and pseudovirus-based neutralization assays. These assays represent the diversity of tests employed for serodiagnosis, serosurveillance, and characterization of levels of binding and neutralizing activity for assessment of efficacy of passive immunotherapies (CCP, monoclonal antibodies, hyperimmune IgG preparations), estimation of vaccine efficacy, and durability of humoral immunity following infection or vaccination that may correlate with protection from reinfections following convalescence or vaccination.

We observed that SARS-CoV-2 antibody assays provided different longitudinal profiles for antibody reactivity over 18 weeks post resolution of COVID-19 symptoms. The commercial direct detection (antigen sandwich format) assays show steady to increasing signals over time while indirect detection IgG assays show declining signals over time. The neutralizing Ab assays had declining responses over time although the change in the BROAD PRNT live virus assay over the 129-day period was not different than the indirect-detection binding assays, whereas the VRI RVPN pseudovirus assay was the most sensitive to the change in viral neutralization over this period.

These findings complement and extend reports from other studies of antibody persistence following infection. The majority of studies have focused on durability of antibodies detected by commercial or lab-developed indirect IgG assays and neutralizing assays, with most studies documenting waning of antibodies following peak reactivity approximately 1 month post-seroconversion.^3,4,5,6,20^ Multiple studies have also correlated the waning of binding assay reactivities, and particularly IgG assays with S1, RBD and NC antigens, relative to waning neutralizing antibodies to identify high throughput and low cost assays that could serve as proxies for live virus PRNT or pseudovirus RVPNT assays.^13,21^ These analyses have demonstrated variable correlations and predictive values of S1, RBD and NC-based immunoassays with waning neutralizing activity. The first high throughput test designated by the US FDA for use in labeling CCP with high- or low-titer antibody content was the Ortho CoV2G IgG assay. Notably, the Abbott Architect SARS-CoV-2 IgG and Ortho Vitros Anti-SARS-CoV-2 IgG assays used in the present study, were among those found acceptable for this purpose.

For serosurveillance studies, which have been widely implemented regionally, nationally and internationally to monitor SARS-CoV-2 transmission dynamics using serial cross-sectional sample sets from populations including blood donors, the preferred assay(s) would have sustained antibody reactivity over at least six months and optimally longer to accurately track cumulative incidence.^19,22,23^ If assays that are susceptible to rapid waning of seroreactivity are employed in serosurveillance studies, significant proportions of previously infected persons could have seroreverted in downstream waves of sample collection and testing.^24^ Such waning can be addressed by statistical adjustments in analyses, based on patterns of waning using CCP sample sets of findings from serial cross-sectional results, as recently done in a large study from Brazil based on the Abbott NC IgG assay included in the present study.^25^ But these adjustments require parallel data on waning profiles, are complex, and have led to debate over the validity and generalizability of serosurveillance findings in studies employing assays that demonstrate antibody waning. In contrast, the direct antigen sandwich assays that we evaluated, which included the S1-based Total Ig assay from Ortho and NC-based Total Ig assay from Roche, are optimal for application in serosurveillance studies given the stability and even increasing levels of reactivity observed over time, presumably due to continued maturation of antibody affinity and/or avidity resulting in increasing signal intensity in these assays.^26,27^ The pattern of persistence and even increasing reactivity of these assays which we observed with serial convalescent plasma donations has also been observed in analyses of serial donations by regular blood donors screened with these assays and by another antigen sandwich configuration assay by Wantai.^28^ Moreover, by combining S- and NC-based direct antigen sandwich assays into algorithms for confirmation of seroreactivity it is possible to minimize the potential contribution of false positive results to estimates of cumulative incidence of natural infections over time and across regional and demographic subgroups. This algorithm has been adopted in the US REDS-IV-P RESPONSE and National Blood Donor Serosurveillance Studies (MASS-BD).

As SARS-CoV-2 vaccines are being rapidly approved and implemented to mitigate the pandemic, there will be a need for combinations of S- and NC-based assays or multiplexed S/RBD/NC assays to detect and discriminate antibodies induced by natural infection from vaccine induced seropositivity (VISP). Assays/algorithms will also be needed to monitor vaccine penetrance and persistence of VISP, as well as to detect breakthrough infections in vaccinated persons following ongoing exposures to SARS-CoV-2. The algorithm adopted for the US National Blood Donor Serosurveillance Study (MASS-BD) that employs the Ortho CoV2T Total Ig assay followed by the Roche NC Total Ig assay on all S1 antibody reactive samples should allow for simultaneous and accurate detection of VISP and natural infection induced seropositivity, and has the potential to surveil for vaccine breakthrough infections in longitudinal databases of repeat blood donors who were determined to be previously vaccinated.

Measuring antibody levels over time within populations, including CCP and routine blood donors, may also provide valuable data on risk of reinfections. Reinfections will likely induce anamnestic boosting of antibody reactivity to both S and NC antigens.^29^ Such boosting will be particularly apparent with assays that are prone to waning, such as the Abbott NC and RBD IgG assays and pseudovirus RVPNT assay. Hence different assays and different combinations of SARS-CoV-2 antibody assays will be needed to address the multiple important questions that are arising as the pandemic and mitigation strategies evolve.

## Data Availability

Data are available by request.

## Acknowledgements

Abbott Diagnostics & Ortho-Clinical Diagnostics kindly provided reagents and testing services for their assay platforms. Thanks and acknowledgements also are extended to the extended teams at Vitalant Research Institute and the Broad Institute that managed sample procurement, preparation, testing, and data verification.

## References

1. Long Q-X, Liu B-Z, Deng H-J, et al. Antibody responses to SARS-CoV-2 in patients with COVID-19. Nat. Med. 2020;26(6):845–848.

2. Cervia C, Nilsson J, Zurbuchen Y, et al. Systemic and mucosal antibody responses specific to SARS-CoV-2 during mild versus severe COVID-19. J. Allergy Clin. Immunol. 2020;S0091674920316237.

3. Long Q-X, Tang X-J, Shi Q-L, et al. Clinical and immunological assessment of asymptomatic SARS-CoV-2 infections. Nat. Med. 2020;26(8):1200–1204.

4. Ibarrondo FJ, Fulcher JA, Goodman-Meza D, et al. Rapid Decay of Anti-SARS-CoV-2 Antibodies in Persons with Mild Covid-19. N. Engl. J. Med. 2020;383(11):1085–1087.

5. Patel MM, Thornburg NJ, Stubblefield WB, et al. Change in Antibodies to SARS-CoV-2 Over 60 Days Among Health Care Personnel in Nashville, Tennessee. JAMA. 2020;324(17):1781.

6. Perreault J, Tremblay T, Fournier M-J, et al. Waning of SARS-CoV-2 RBD antibodies in longitudinal convalescent plasma samples within 4 months after symptom onset. Blood. 2020;136(22):2588–2591.

7. Wajnberg A, Amanat F, Firpo A, et al. Robust neutralizing antibodies to SARS-CoV-2 infection persist for months. Science. 2020;370(6521):1227–1230.

8. Dan JM, Mateus J, Kato Y, et al. Immunological memory to SARS-CoV-2 assessed for up to 8 months after infection. Science. 2021;eabf4063.

9. Choe PG, Kang CK, Suh HJ, et al. Waning Antibody Responses in Asymptomatic and Symptomatic SARS-CoV-2 Infection - Volume 27, Number 1—January 2021 - Emerging Infectious Diseases journal - CDC.

10. Kellam P, Barclay W. The dynamics of humoral immune responses following SARS-CoV-2 infection and the potential for reinfection. J. Gen. Virol. 2020;101(8):791–797.

11. U.S. Food and Drug Administration. EUA Authorized Serology Test Performance. 2021;

12. U.S. Food and Drug Administration. Investigational COVID-19 Convalescent Plasma; Guidance for Industry. 2021;

13. Goodhue Meyer E, Simmons G, Grebe E, et al. Selecting COVID-19 convalescent plasma for neutralizing antibody potency using a high-capacity SARS-CoV-2 antibody assay. Transfusion (Paris). 2021;10.1111/trf.16321.

14. Ortho Clinical Diagnostics. VITROS Immunodiagnostic Products Anti-SARS-CoV-2 Total Reagent Pack and VITROS Immunodiagnostic Products Anti-SARS-CoV-2 Total Calibrator Instructions for Use. Version 3.0. Ortho Clinical Diagnostics, Inc. Rochester, New York. April 14, 2020. https://www.orthoclinicaldiagnostics.com/. 2020;

15. Elecsys Anti-SARS-CoV-2 S. Instructions for Use Version 1.0. Roche Diagnostics GmbH, Sandhofer Strasse 116, D-68305 Mannheim. 12/2020. https://diagnostics.roche.com/us/en/products/params/elecsys-anti-sars-cov-2-s.html. 2020;

16. Ortho Clinical Diagnostics. VITROS Immunodiagnostic Products Anti-SARS-CoV-2 IgG Reagent Pack and VITROS Immunodiagnostic Products Anti-SARS-CoV-2 IgG Calibrator Instructions for Use. Version 4.2. Ortho Clinical Diagnostics, Inc. Rochester, New York. April 14, 2020.https://www.orthoclinicaldiagnostics.com/. 2020;

17. Architect SARS-CoV-2 IgG Instructions for Use. G90418R01. Abbott Laboratories Diagnostics Division Abbott Park, IL 60064 USA. https://www.abbott.com/for-healthcare-professionals/diagnostics.html. 2020;

18. Hoffmann M, Kleine-Weber H, Schroeder S, et al. SARS-CoV-2 Cell Entry Depends on ACE2 and TMPRSS2 and Is Blocked by a Clinically Proven Protease Inhibitor. Cell. 2020;181(2):271-280.e8.

19. Ng DL, Goldgof GM, Shy BR, et al. SARS-CoV-2 seroprevalence and neutralizing activity in donor and patient blood. Nat. Commun. 2020;11(1):4698.

20. Peluso MJ, Takahashi S, Hakim J, et al. SARS-CoV-2 antibody magnitude and detectability are driven by disease severity, timing, and assay. medRxiv. 2021;2021.03.03.21251639.

21. Salazar E, Kuchipudi SV, Christensen PA, et al. Convalescent plasma anti–SARS-CoV-2 spike protein ectodomain and receptor-binding domain IgG correlate with virus neutralization. J. Clin. Invest. 2020;130(12):6728–6738.

22. Havers FP, Reed C, Lim T, et al. Seroprevalence of Antibodies to SARS-CoV-2 in 10 Sites in the United States, March 23-May 12, 2020. JAMA Intern. Med. 2020;180(12):1576.

23. Rosenberg ES, Tesoriero JM, Rosenthal EM, et al. Cumulative incidence and diagnosis of SARS-CoV-2 infection in New York. Ann. Epidemiol. 2020;48:23–29.e4.

24. Shioda K, Lau MS, Kraay AN, et al. Estimating the cumulative incidence of SARS-CoV-2 infection and the infection fatality ratio in light of waning antibodies. medRxiv. 2020;10.1101/2020.11.13.20231266:

25. Buss LF, Prete CA, Abrahim CMM, et al. Three-quarters attack rate of SARS-CoV-2 in the Brazilian Amazon during a largely unmitigated epidemic. Science. 2021;371(6526):288–292.

26. Liu H, Wu NC, Yuan M, et al. Cross-Neutralization of a SARS-CoV-2 Antibody to a Functionally Conserved Site Is Mediated by Avidity. Immunity. 2020;53(6):1272–1280.e5.

27. Benner SE, Patel EU, Laeyendecker O, et al. SARS-CoV-2 Antibody Avidity Responses in COVID-19 Patients and Convalescent Plasma Donors. J. Infect. Dis. 2020;222(12):1974–1984.

28. GeurtsvanKessel CH, Okba NMA, Igloi Z, et al. An evaluation of COVID-19 serological assays informs future diagnostics and exposure assessment. Nat. Commun. 2020;11(1):3436.

29. Babiker A, Marvil C, Waggoner JJ, Collins M, Piantadosi A. The Importance and Challenges of Identifying SARS-CoV-2 Reinfections. J. Clin. Microbiol. 2020;10.1128/JCM.02769-20:.

